# Does Medication Order Matter? Incident Dementia Risk When Gabapentin Is Added to Prior Dihydropyridine Calcium Channel Blocker Therapy: An Active-Comparator Cohort Study in Hypertensive Adults

**DOI:** 10.64898/2026.03.30.26349801

**Authors:** James W. Green, Joshua Kaplan, Branimir Ljubic, Michael Schulewski, Suril Gohel, Sharon Sanz Simon, Luciana Mascarenhas Fonseca, Michal Schnaider Beeri, Laura Byham-Gray, Barbara Tafuto

## Abstract

**Background:** Older adults frequently take both gabapentinoids and dihydropyridine calcium channel blockers (DHP-CCBs), often started years apart as new conditions emerge. A common pattern is the late addition of gabapentin to a regimen that has long included a DHP-CCB. Prior work links concurrent gabapentinoid–DHP-CCB exposure to elevated incident dementia risk, but whether the order in which the two medications are introduced matters has not been examined. **We asked whether incident dementia risk differs by the order in which gabapentinoid and DHP-CCB therapy are introduced**.

**Methods:** Using the Rutgers Clinical Research Data Warehouse (2018–2024) and active-comparator new-user designs with IPTW Cox models, we examined two prescribing scenarios in a hypertensive cohort: gabapentin or pregabalin added after prior DHP-CCB exposure (forward sequence; N=7,141), and a DHP-CCB or ACE inhibitor/ARB added after chronic gabapentinoid therapy (reverse sequence; N=685).

**Results:** When gabapentin was added to a regimen that already included a DHP-CCB, incident dementia risk was elevated relative to pregabalin (**IPTW HR 2.44, 95% CI 1.06–5.63, p=0.036**). When the order was reversed — a DHP-CCB added to chronic gabapentinoid therapy — no elevated risk was observed (**IPTW HR 0.64, 95% CI 0.17– 2.39**). Pre-specified sensitivity analyses supported the forward-sequence finding.

**Conclusions:** In hypertensive older adults, the same two medications were associated with very different dementia signals depending on which was started first: elevated risk when gabapentin was added to chronic DHP-CCB therapy, no elevated signal in the reverse direction. The two prescribing sequences may not be clinically equivalent. If replicated, this pattern would support targeted medication review at the time gabapentin is added to regimens that already include a DHP-CCB.

## Introduction

Older adults rarely begin a complex medication regimen all at once. They typically transition onto chronic medications sequentially over years as new conditions emerge, and the order in which medications are introduced is determined more by the timing of new diagnoses than by any single prescribing plan. Among the most common patterns in this population is the late addition of gabapentin to a regimen that already includes long-standing antihypertensive therapy: a patient managed for years on amlodipine for hypertension develops painful peripheral neuropathy at age 70 and is started on gabapentin. Gabapentinoids—gabapentin and pregabalin—rank among the most widely prescribed medications in the United States, with gabapentin dispensed to approximately 15.5 million patients annually,^1^ predominantly to older adults who frequently carry concurrent cardiovascular diagnoses and who, by the time gabapentin is initiated, have often been on a dihydropyridine calcium channel blocker (DHP-CCB) for years.^2^ Despite this typical clinical pattern, no study has examined whether the order in which gabapentinoids and DHP-CCBs are introduced influences subsequent cognitive outcomes.

Prior pharmacoepidemiologic studies have reported associations between gabapentinoid use and incident dementia or cognitive decline, with mixed findings. A large U.S. EHR cohort study found a 29% elevated dementia risk with chronic gabapentin use in adults with chronic pain (RR 1.29, 95% CI 1.18–1.40), with a dose-response relationship by prescription frequency.^3^ A retrospective study using the National Alzheimer’s Coordinating Center dataset reported cognitive and functional decline following gabapentin initiation in older adults with normal cognition.□ A population-based cohort of more than 200,000 patients in Taiwan similarly found increased dementia risk with gabapentinoid use.□ A nested case-control analysis of 201,492 chronic-pain patients found no significant association (adjusted OR 0.91, 95% CI 0.83–1.01), highlighting the inconsistency in this literature.□ None of these studies stratified by concurrent calcium channel blocker exposure. In related work from our research program, DHP-CCB co-medication was associated with substantially elevated gabapentin-dementia risk (HR 2.22 versus HR 1.15 in non-CCB users), with subtype specificity consistent with neuronal L-type calcium channel selectivity as one plausible mechanism.□ A companion analysis from the same program reported partial reversibility of acute cognitive effects upon discontinuation of the combination.□ Whether the gabapentinoid–DHP-CCB signal depends on the order in which the two drugs are initiated has not been examined.

The clinical question is straightforward: does it matter whether gabapentin is added to chronic DHP-CCB therapy versus the reverse? Two patients can end up on the same combination of drugs by different routes — one starts amlodipine first, then gabapentin years later; the other starts gabapentin first, then amlodipine. The pharmacology textbook treats these as equivalent. Clinical experience suggests they may not be: the body adjusts to whichever drug arrives first, and the effect of the second may depend on what was already on board. Whether this matters for incident dementia in patients on both classes is unknown. If sequence does matter, it would have practical implications for when medication review is most informative — at gabapentin initiation in CCB-treated patients, at antihypertensive initiation in gabapentin-treated patients, or both. Several mechanisms could in principle produce sequence-dependent effects, but observational data cannot distinguish among them; we treat such hypotheses as exploratory only.

### Objective

We asked whether the order in which gabapentinoid and DHP-CCB therapy are introduced is associated with differential incident dementia risk in hypertensive adults. Specifically, we compared two clinical scenarios: (1) gabapentin or pregabalin added to a regimen that already included a DHP-CCB (the forward sequence), and (2) a DHP-CCB or ACE inhibitor/ARB added to a regimen that already included chronic gabapentinoid therapy (the reverse sequence). We used an active-comparator new-user design across three pre-specified cohorts: a primary cohort of gabapentinoid initiators with any prior DHP-CCB exposure (Broader CCB-first), a stricter chronic-CCB sensitivity cohort (Pop 4), and a reverse-sequence cohort (Pop 3). This is, to our knowledge, the first pharmacoepidemiologic study to examine whether prescribing order modifies the gabapentinoid–DHP-CCB dementia association. We frame the analysis as a focused clinical timing study, not as a test of any specific biological mechanism.

## Methods

### Data Source

We used a hypertension-focused research extract from the Rutgers Clinical Research Data Warehouse (CRDW)^11^, comprising 541,539 patients with documented encounter, medication, and comorbidity data from 2018–2024, serving the Rutgers University health system in northern New Jersey. The Rutgers Institutional Review Board approved this study with waiver of informed consent.

### Medication Exposure Ascertainment

Medication exposure was identified from the CRDW prescription file (Hypertension Medications 20250417) using a generic-name–first matching strategy. For each prescription record, we matched on the standardized generic name (SIMPLE_GENERIC) when present, falling back to the full medication name (MEDICATION_NAME) when SIMPLE_GENERIC was missing. To restrict the analysis to sustained outpatient oral therapy, we excluded prescription records with ACTIVE_ORDER_STATUS = ‘Discontinued Medication’ and excluded intravenous formulations identified by route (MED_ADMIN_ROUTE containing ‘intravenous’, ‘injection’, ‘iv push’, ‘iv bolus’, or ‘iv drip’) or by product-name pattern (e.g., ‘…intravenous solution’, ‘…in 0.9% sodium chloride’, ‘…mg/mL intravenous’). Drug-class definitions (DHP-CCB, non-DHP-CCB, ACE-inhibitor, ARB) used the standardized generic-name list specified in the supplementary methods. Ascertainment was performed at the prescription-record level prior to cohort-level eligibility filtering.

### Master Cohort and Sequence Classification

The master cohort comprised 33,791 hypertensive patients with at least one gabapentinoid prescription during the observation window. We classified patients by the relative timing of first DHP-CCB and first gabapentinoid prescription using corrected medication ascertainment. Sequence categories were: CCB-first (DHP-CCB before gabapentinoid index, N=7,033, 20.8%); Same-day (N=4,551, 13.5%); Gabapentinoid-first (gabapentinoid ≥30 days before any DHP-CCB, N=2,729, 8.1%); No-CCB (N=17,344, 51.3%); and Unknown (gabapentinoid temporal anchor not establishable under corrected ascertainment, N=2,134, 6.3%). The Unknown category increased modestly relative to prior versions of this work because the corrected pipeline excludes patients whose only gabapentinoid records were discontinued or intravenous; this is a methodologically conservative filter and the affected patients are excluded from all analyses.

### Primary Analysis: Broader CCB-First Cohort

The primary analysis applied an active-comparator new-user design comparing gabapentin versus pregabalin in the Broader CCB-first cohort: gabapentinoid initiators with any prior DHP-CCB prescription before gabapentinoid index, with no prior gabapentinoid exposure within 12 months and ≥180 days of prior observation. We excluded patients with pre-existing dementia diagnoses. After dropping records with missing exposure, event, or follow-up time, the analytic dataset comprised 7,141 patients (gabapentin N=5,994; pregabalin N=1,147) with 82 incident dementia events. The exposure was gabapentin versus pregabalin (active-comparator design); the primary outcome was incident all-cause dementia (ICD-10 codes F01–F03, F00.x, G30, G31.0, G31.01, G31.09, G31.1, G31.83, G31.85) following gabapentinoid index date. Follow-up continued until dementia diagnosis, death, last healthcare encounter, or December 31, 2024. Statistical methods are detailed in the consolidated Statistical Analysis subsection below.

### Recency Sensitivity Analysis

To examine whether the temporal relationship between prior CCB exposure and gabapentinoid initiation modifies the effect estimate, we conducted recency-restricted analyses in the Broader CCB-first cohort, restricting prior DHP-CCB exposure to within 365, 180, 90, and 30 days of gabapentinoid index. We pre-specified 365 days as the principal recency window of interest (corresponding to a clinically meaningful “current or recent” exposure). Narrower windows are reported as exploratory sensitivity analyses; we do not interpret these as evidence for a monotonic recency dose-response.

### Stricter Sensitivity Analysis: Pop 4 (Chronic CCB)

As a stricter sensitivity analysis, we restricted the prior DHP-CCB criterion to chronic CCB use defined as ≥2 fills spanning ≥90 days before gabapentinoid initiation (Pop 4; analytic N=2,592 after corrected ascertainment; 25 events).

### Lag Sensitivity Analyses (Protopathic-Bias Assessment)

To assess potential protopathic bias—the concern that gabapentinoid initiation may have been prompted by emerging cognitive symptoms not yet diagnosed at index—we conducted lagged sensitivity analyses excluding incident dementia events occurring within 30, 60, and 90 days of gabapentinoid index in the Broader CCB-first cohort. Protopathic bias predicts that hazard ratios should attenuate as early events are excluded; persistence or strengthening of the effect estimate across lag conditions provides reassurance against this bias.

### Sensitivity to Unmeasured Confounding (E-value)

We computed the VanderWeele-Ding E-value as a sensitivity-to-confounding measure for the primary IPTW HR.^12^ The E-value represents the minimum strength of association on the risk-ratio scale that an unmeasured confounder would need to have with both treatment and outcome to fully explain away the observed association. We report E-values for both the point estimate and the lower 95% confidence interval bound.

### Reverse-Sequence Analysis: Population 3

To examine the reverse prescribing order, we identified patients on established chronic gabapentinoid therapy (≥2 fills spanning ≥90 days) who newly initiated an antihypertensive—either a DHP-CCB or an ACE inhibitor/ARB—with no prior prescription of the comparator class within 12 months. The contrast was DHP-CCB versus ACE/ARB initiation, an active comparator within the reverse sequence.

### Established Dementia Cohort (Exploratory)

As an exploratory analysis conceptually separate from the primary incident-dementia analyses, we identified patients with established dementia (first diagnosis ≥180 days before gabapentinoid initiation) who newly initiated gabapentinoid therapy, classified by CCB status at initiation. An IPTW Cox model estimated the hazard ratio for incident encephalopathy comparing patients with chronic outpatient DHP-CCB exposure to those with no CCB exposure; the non-DHP CCB arm is reported descriptively given small N. This analysis reflects acute adverse-event risk in a high-risk population and should not be interpreted as evidence of accelerated neurodegeneration. Results are reported in Supplemental Table S1.

### Statistical Analysis

All hazard-ratio estimates used inverse probability of treatment weighting (IPTW) Cox proportional hazards models. Propensity scores for treatment assignment (gabapentin vs. pregabalin in the primary, recency, and chronic-CCB analyses; DHP-CCB vs. ACE inhibitor/ARB in the reverse-sequence analysis; DHP-CCB vs. no CCB in the established dementia exploratory cohort) were estimated with L2-penalized logistic regression (ridge; C=1.0) on age, sex, and baseline CKD, with gabapentinoid duration before index added in Pop 3 and dementia duration before index added in the established dementia cohort.^13^ IPTW used average-treatment-effect-on-the-treated (ATT) weights, truncated at the 1st and 99th percentiles and stabilized; covariate balance was assessed by standardized mean differences with a target threshold of post-weighting |SMD| <0.10. Pre-specified sensitivity analyses included the recency-restricted, chronic-CCB, lagged, reverse-sequence, and E-value analyses described above. Two-sided p<0.05 was the threshold for statistical significance throughout. Statistical analyses were conducted in Python (lifelines for Cox models; scikit-learn for propensity-score estimation).

### Pre-Specification and Reporting

The primary pre-specified contrast was gabapentin versus pregabalin initiation in the Broader CCB-first cohort. Population 3 (DHP-CCB vs ACE/ARB in chronic gabapentinoid users) was pre-specified as the reverse-sequence analysis. Pop 4 (chronic-CCB criterion) was pre-specified as a stricter sensitivity analysis. The 365-day recency cut was the pre-specified principal recency window. Reporting follows STROBE guidelines for observational pharmacoepidemiology.^1^ □;

## Results

We organize the Results around the prescribing-order question. The forward-sequence analysis (gabapentin or pregabalin added to prior DHP-CCB therapy) is presented first, followed by the reverse-sequence analysis (DHP-CCB or ACE/ARB added to chronic gabapentinoid therapy), and finally the supporting sensitivity analyses.

### Cohort Characteristics

Table 1 presents baseline characteristics across the analytic cohorts. The Broader CCB-first analytic cohort (N=7,141) had a mean age of approximately 69.5 years, with the majority female; 80%–82% initiated gabapentin and 18%–20% initiated pregabalin. The stricter Pop 4 chronic-CCB sensitivity cohort (N=2,592) showed a similar age and sex distribution. Population 3 comprised patients on chronic gabapentinoid therapy initiating a new antihypertensive (DHP-CCB or ACE/ARB). IPTW achieved acceptable covariate balance in all three cohorts (post-weighting |SMD| <0.10 in the primary and Pop 4 cohorts; |SMD| <0.01 in Pop 3).

**Table 1.**
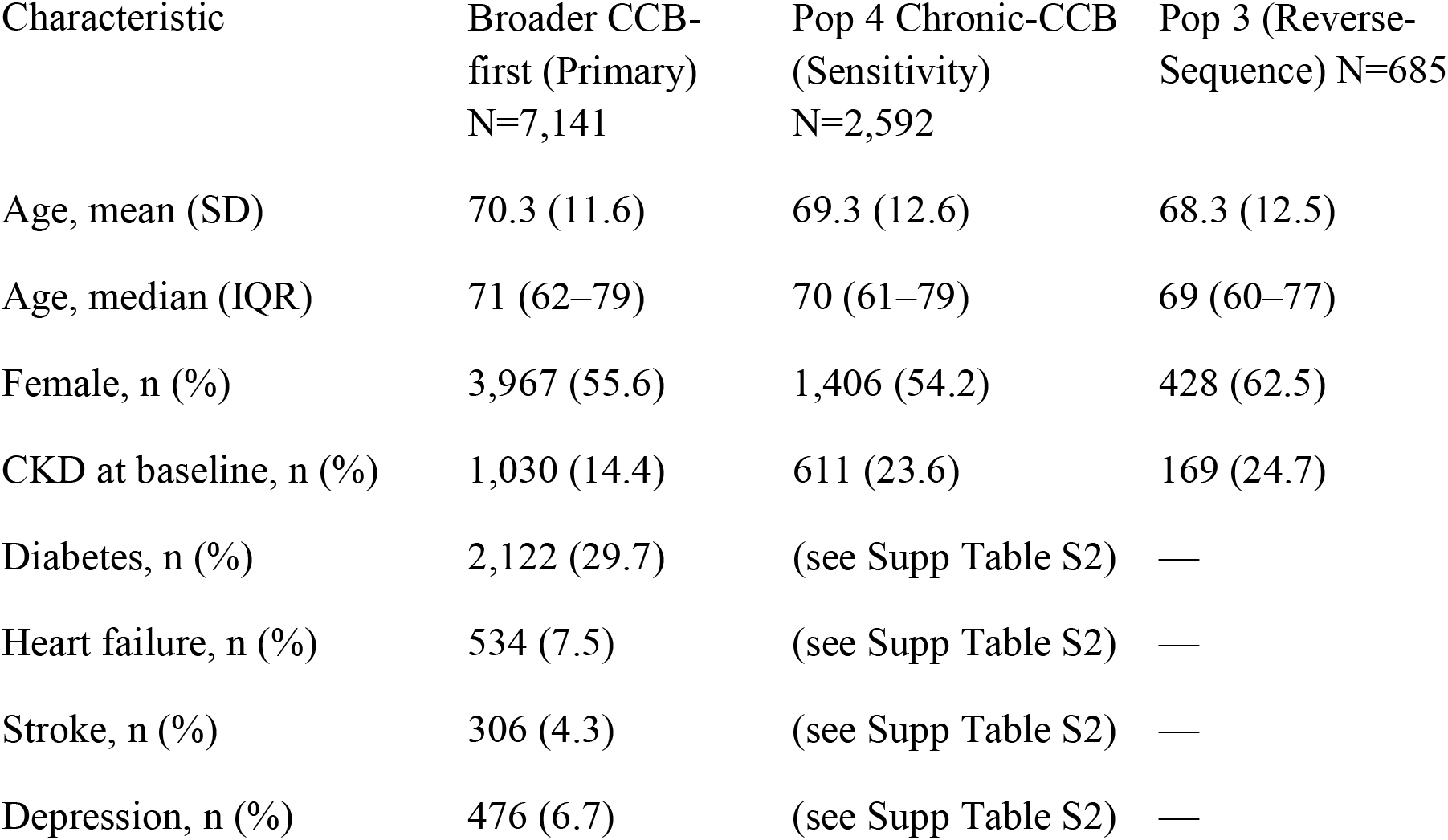
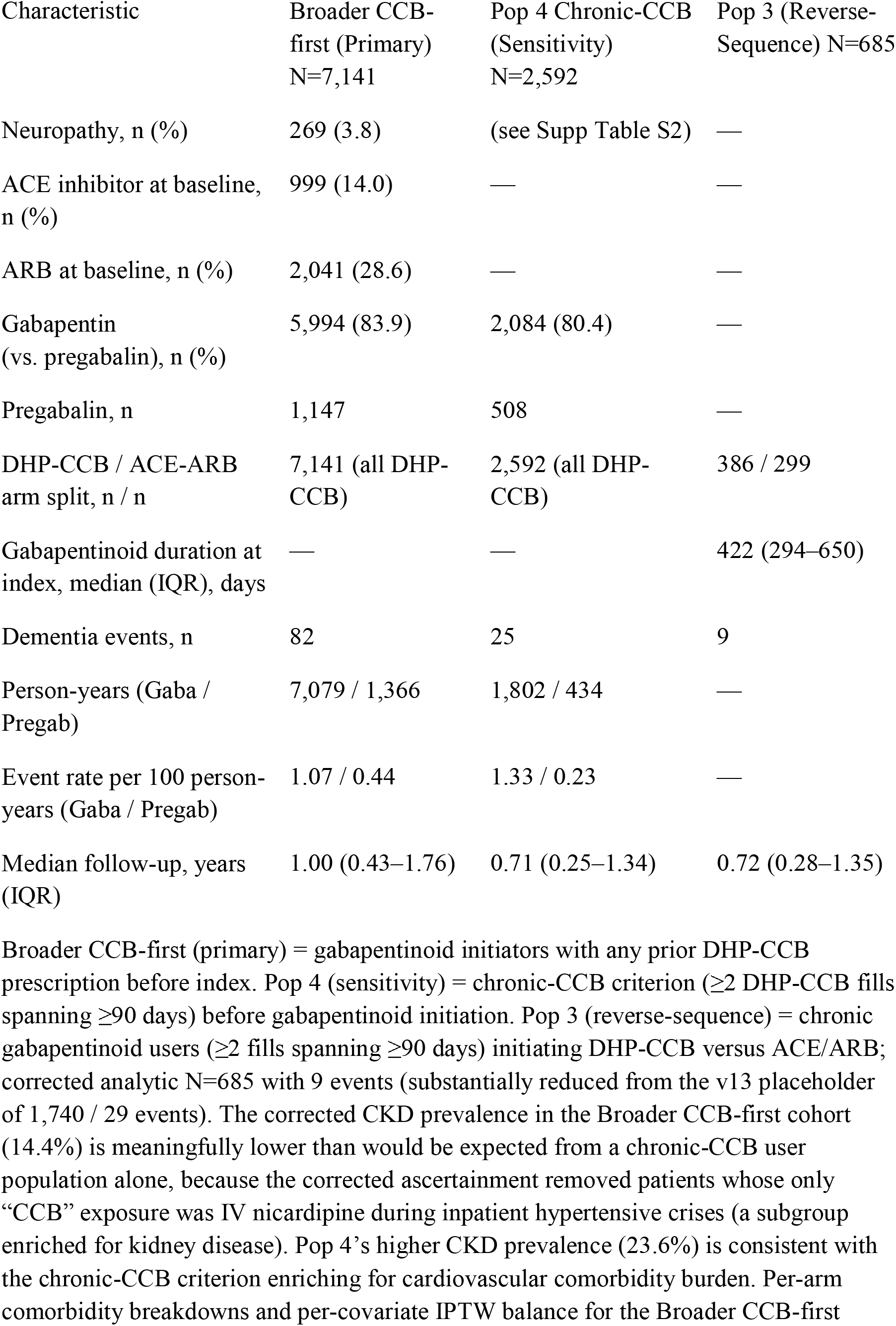
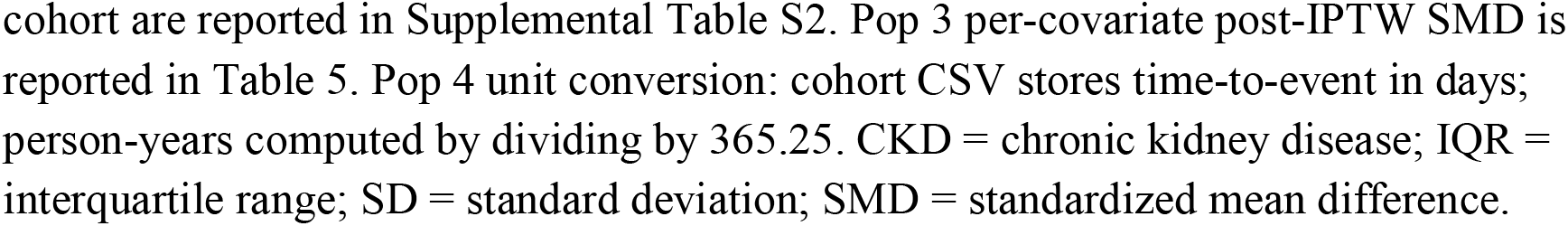
Cohort Characteristics by Analytic Population.

### Primary Analysis: Broader CCB-First Cohort

Among hypertensive adults with prior DHP-CCB exposure, dementia incidence was higher among those who started gabapentin than among those who started pregabalin (1.07 vs. 0.44 per 100 person-years; 76 vs. 6 events). The corresponding IPTW Cox hazard ratio was 2.44 (95% CI 1.06–5.63, p=0.036; Table 2; E-value 4.31 for the point estimate, 1.31 for the lower CI bound).

**Table 2.** Primary Analysis: Gabapentin versus Pregabalin in the Broader CCB-First Cohort.

| Comparison | N | Events | Person-years | Rate per 100 person-years | Crude HR (95% CI) | IPTW HR (95% CI) | P-value |
| --- | --- | --- | --- | --- | --- | --- | --- |
| Gabapentin vs. pregabalin (any prior DHP-CCB) | 7,141 | 82 | 8,445 | — | 2.45 (1.07–5.63) | 2.44 (1.06–5.63) | 0.036 |
| Gabapentin arm | 5,994 | 76 | 7,079 | 1.07 | — | Reference: elevated | — |
| Pregabalin arm | 1,147 | 6 | 1,366 | 0.44 | — | Reference: lower | — |

Primary pre-specified analysis. Population: gabapentinoid initiators with any prior DHP-CCB prescription before index, $\geq 180$ days of prior observation, no prior gabapentinoid within 12 months, no pre-existing dementia. IPTW used L2-penalized logistic regression ( $C=1.0$ ) with stabilized ATT weights truncated at the 1st/99th percentiles; all post-IPTW $|SMD| < 0.10$ . VanderWeele-Ding E-value: 4.31 for the point estimate, 1.31 for the lower 95% CI bound. HR = hazard ratio; IPTW = inverse probability of treatment weighting.

### Reverse-Sequence Analysis: Population 3

When the prescribing order was reversed — patients on chronic gabapentinoid therapy who newly added a DHP-CCB versus an ACE inhibitor or ARB — no elevated dementia risk was observed (IPTW HR 0.64, 95% CI 0.17–2.39, p=0.51; 9 events in 685 patients; Table 5). The wide CI reflects event sparsity; we read this as a non-elevated finding consistent with the directional clinical observation.

### Recency Sensitivity Analysis

The signal was concentrated in patients whose prior DHP-CCB exposure was clinically recent: restricting prior CCB exposure to within 365 days of gabapentinoid initiation strengthened the effect estimate (IPTW HR 3.65, 95% CI 1.14–11.65; Table 3). Narrower windows did not preserve statistical significance, reflecting event sparsity in the pregabalin arm. We do not interpret these data as evidence of a monotonic recency gradient.

**Table 3.** Recency Sensitivity in the Broader CCB-First Cohort.

| Prior DHP-CCB Exposure Window | Total N | Events | Gaba N / Events | Pregab N / Events | IPTW HR (95% CI) | P-value |
| --- | --- | --- | --- | --- | --- | --- |
| **Any prior DHP-CCB (primary)** | **7,141** | **82** | **5,994 / 76** | **1,147 / 6** | **2.44 (1.06–5.63)** | **0.036** |
| **$\leq 365$ days before index (pre-specified recency cut)** | **3,735** | **59** | **3,123 / 56** | **612 / 3** | **3.65 (1.14–11.65)** | **0.029** |
| $\leq 180$ days before index (exploratory) | 2,125 | 27 | 1,752 / 24 | 373 / 3 | 1.66 (0.50–5.49) | 0.41 |
| ≤90 days before index (exploratory) | 1,388 | 16 | 1,145 / 14 | 243 / 2 | 1.45 (0.33–6.38) | 0.62 |
| ≤30 days before index (exploratory) | 716 | 6 | 598 / 6 | 118 / 0 | Not estimable | — |

Bolded rows are the pre-specified primary analysis and the pre-specified principal recency window; lower rows are exploratory. The ≤30-day window was not estimable because the pregabalin arm contained zero events at that restriction; the ≤180-day and ≤90-day windows had 3 and 2 pregabalin events respectively, accounting for the wide intervals at narrow windows.

### Stricter Sensitivity Analysis: Pop 4 (Chronic CCB)

Restricting to chronic CCB use (≥2 fills spanning ≥90 days) preserved direction with reduced precision (IPTW HR 4.81, 95% CI 0.66–35.05; Table 4), reflecting event sparsity in the pregabalin arm.

**Table 4.** Stricter Sensitivity Analysis: Pop 4 Chronic-CCB Criterion.

| Comparison | N | Events | Person-years | Rate per 100 py | IPTW HR (95% CI) | P-value |
| --- | --- | --- | --- | --- | --- | --- |
| Gabapentin vs. pregabalin (chronic CCB ≥2 fills, ≥90 d) | 2,592 | 25 | 2,236 | 1.12 | 4.81 (0.66–35.05) | 0.12 |
| Gabapentin arm | 2,084 | 24 | 1,802 | 1.33 | — | — |
| Pregabalin arm | 508 | 1 | 434 | 0.23 | — | — |

Stricter sensitivity analysis. Chronic CCB defined as ≥2 DHP-CCB fills spanning ≥90 days before gabapentinoid initiation. Person-years computed by dividing time-to-event in days by 365.25. The wide confidence interval reflects event sparsity in the pregabalin arm (1 event in 508 initiators).

**Table 5.** Reverse-Sequence Analysis: Population 3 (Chronic Gabapentinoid → Add DHP-CCB vs. ACE/ARB)

| Comparison | N | Events | Crude HR (95% CI) | IPTW HR (95% CI) | P-value |
| --- | --- | --- | --- | --- | --- |
| DHP-CCB vs. ACE/ARB (chronic gabapentinoid users) | 685 | 9 | 0.70 (0.19–2.60) | 0.64 (0.17–2.39) | 0.51 |

**Pop 3 per-covariate post-IPTW standardized mean differences:**
| Covariate | Pre-IPTW<br>SMD | Post-IPTW<br>SMD | Status |
| --- | --- | --- | --- |
| Age (continuous) | +0.121 | −0.002 | balanced<br>□ |
| Age ≥65 (binary) | +0.165 | −0.000 | balanced<br>□ |
| Sex (male) | −0.047 | −0.001 | balanced<br>□ |
| CKD at baseline | +0.163 | +0.000 | balanced<br>□ |
| Gabapentinoid duration before index | +0.085 | −0.000 | balanced<br>□ |

Pre-specified reverse-sequence analysis. Population: chronic gabapentinoid users ( $\geq 2$ fills spanning $\geq 90$ days) who newly initiated either a DHP-CCB or an ACE inhibitor/ARB, with no prior prescription of the comparator class within 12 months. Contrast: DHP-CCB versus ACE/ARB. The wide confidence interval reflects the small event count (9 events in 685 patients). All post-IPTW $|SMD| < 0.10$ .

The remaining results present three pre-specified sensitivity analyses that support the forward-sequence finding.

### Lag Sensitivity Analyses (Protopathic-Bias Assessment)

The signal did not attenuate when events soon after gabapentinoid initiation were excluded — arguing against protopathic bias. Lagged analyses (excluding events within 30, 60, or 90 days of index) produced IPTW HRs that all exceeded the no-lag estimate (Table 6, all p<0.05).

**Table 6.** Protopathic-Bias Assessment: Lagged Sensitivity Analyses in the Broader CCB-First Cohort.

| Lag Window | N | Events | IPTW HR (95% CI) | P-value | $\Delta$ from lag-0 |
| --- | --- | --- | --- | --- | --- |
| **No lag (primary)** | **7,141** | **82** | **2.44 (1.06–5.63)** | **0.036** | **—** |
| 30-day lag (exclude events $\leq 30$ d) | 6,699 | 76 | 2.88 (1.16–7.13) | 0.023 | +17.7% |
| 60-day lag (exclude events $\leq 60$ d) | 6,337 | 71 | 2.74 (1.10–6.80) | 0.030 | +12.1% |
| 90-day lag (exclude events $\leq 90$ d) | 6,027 | 64 | 3.15 (1.14–8.70) | 0.027 | +29.0% |

Protopathic-bias assessment by excluding incident dementia events within 30, 60, and 90 days of gabapentinoid index. Protopathic bias would predict attenuation of HRs as early events are excluded; the persistence and modest strengthening across lag conditions is inconsistent with this bias.

### Established Dementia Cohort (Exploratory)

In the exploratory established-dementia cohort under the strengthened ascertainment, DHP-CCB exposure at gabapentinoid initiation was not associated with differential incident encephalopathy risk relative to no CCB exposure (IPTW HR 0.73, 95% CI 0.30– 1.76, p=0.48; Supplemental Table S1; 8 events of 101 DHP-CCB-exposed [7.9%] vs. 15 events of 145 no-CCB [10.3%]). The descriptive non-DHP-CCB arm (N=8, 0 events) was too small to inform subtype-specificity comparisons. The corrected null contrasts with an uncorrected analysis (HR 3.18, 95% CI 1.36–7.46) that was inflated by intravenous nicardipine prescriptions captured during inpatient hypertensive emergencies; the corrected analysis restricts to chronic outpatient DHP-CCB exposure and removes this source of bias. These results reflect acute adverse-event risk in a high-risk population and should not be interpreted as evidence of accelerated neurodegeneration.

## Discussion

In this single-system EHR study, the clinical order in which gabapentin and DHP-CCBs were introduced was associated with differential incident dementia risk. When gabapentin was added to a regimen that already included a DHP-CCB, dementia incidence was higher than when pregabalin was added in the same setting (HR 2.44; HR 3.65 with recent CCB exposure). When the order was reversed, no elevated risk was observed (HR 0.64). Pre-specified sensitivity analyses supported the forward-sequence finding. These results raise the possibility that the two prescribing sequences are not clinically equivalent, with practical implications for the timing of medication review.

### Directional clinical observation

The clinical observation is the simpler statement: the same two medications may not be clinically equivalent depending on which was started first. Among hypertensive adults with both medications eventually on board, dementia incidence was elevated when gabapentin was added second (after a DHP-CCB) but not when a DHP-CCB was added second (after chronic gabapentinoid therapy). The Pop 3 estimate is imprecise (HR 0.64, 95% CI 0.17–2.39; 9 events), and we do not weight its specific value; the informative comparison is between directions, not between point estimates. Several explanations are plausible, but this study was not designed to distinguish among them.

### Supportive sensitivity analyses

The strengthened estimate when prior DHP-CCB exposure was restricted to within 365 days (HR 3.65) is consistent with recent rather than remote exposure as the relevant clinical condition. A stricter chronic-CCB criterion preserved direction (HR 4.81, 95% CI 0.66–35.05), with magnitude not reliably estimable at this event count. Lagged analyses excluding events within 30, 60, or 90 days of index yielded IPTW HRs of 2.88–3.15, arguing against protopathic bias. The E-value for the primary point estimate was 4.31 (1.31 for the lower CI bound): the direction is robust to modest unmeasured confounding, statistical significance less so.

### Replication and design implications

Replication in event-rich datasets — Medicare fee-for-service in particular — will be necessary to estimate effect magnitude with greater precision and to evaluate the directional observation against larger reverse-sequence cohorts; we are pursuing this via the Rutgers Institute for Health Data Core^1^□. Within the limits of a single-center cohort, these findings support a clinically interpretable directional observation about prescribing order rather than a precise estimate of how much the two sequences differ.

## Limitations

Several limitations warrant discussion. First, we relied on EHR prescription records to ascertain chronic outpatient medication exposure; despite excluding discontinued orders and intravenous formulations, residual misclassification (non-adherence, prescriptions filled outside the system) cannot be excluded. Pharmacoepidemiology studies in this data environment are sensitive to assumptions about which records reflect sustained outpatient use; the exploratory established-dementia analysis (Supplemental Table S1) is illustrative — an apparent positive signal under an unfiltered pipeline (HR 3.18, 95% CI 1.36–7.46) was driven by intravenous nicardipine captured during inpatient hypertensive emergencies and resolved to null (HR 0.73, 95% CI 0.30–1.76) once chronic outpatient exposure was isolated.

Second, the pregabalin comparator arm contained substantially fewer patients and events than the gabapentin arm, producing wider confidence intervals throughout. This is most consequential in the stricter chronic-CCB sensitivity analysis (Pop 4) and the reverse-sequence analysis (Pop 3), where the analytic cohorts shrank substantially under the strengthened ascertainment (Pop 3: 1,740→685 patients, 29→9 events). The directional clinical observation is preserved, but the precision of the Pop 3 estimate is reduced. Third, the pre-specified recency-gradient hypothesis was not confirmed: while the 365-day cut produced a strengthened estimate, narrower windows did not preserve statistical significance. The data are consistent with an exposure window of months to a year as the relevant clinical context; residual confounding by indication and statistical instability at small event counts cannot be excluded.

Fourth, dementia ascertainment relied on EHR-coded diagnoses without neuroimaging confirmation or outcome adjudication; outcome misclassification cannot be excluded. The relatively short median follow-up reflects the available CRDW observation window and limits interpretation of long-latency outcomes. Fifth, all findings derive from a single academic health system in northern New Jersey and may not generalize. Medicare fee-for-service replication via the Rutgers Institute for Health Data Core (DUA RSCH-2024-70249) is in progress.

## Conclusions

In hypertensive older adults, the order in which gabapentin and DHP-CCB therapy were introduced was associated with differential incident dementia risk in this single-system EHR study. When gabapentin was added to a regimen that already included a DHP-CCB, dementia incidence was higher than when pregabalin was added in the same setting (IPTW HR 2.44, 95% CI 1.06–5.63), with a stronger estimate when prior CCB exposure was clinically recent (HR 3.65 within 365 days). When the order was reversed — a DHP-CCB added to chronic gabapentinoid therapy — no elevated risk relative to adding an ACE inhibitor or ARB was observed (HR 0.64, 95% CI 0.17–2.39). Lag-sensitivity analyses argued against protopathic bias. Taken together, the two prescribing sequences may not be clinically equivalent, though precision is limited by the available event count. These findings are hypothesis-generating and require replication in larger event-rich cohorts before informing prescribing guidance. If replicated, they would support targeted medication review at the time gabapentin is added to existing antihypertensive regimens that include a DHP-CCB.

## Data Availability

The Rutgers Clinical Research Data Warehouse (CRDW) data that support the findings of this study are available through the Rutgers University health system subject to institutional data governance review. Analytic code is available from the corresponding author upon reasonable request.

## Acknowledgments

Data used in this research were obtained from the Clinical Research Data Warehouse (CRDW), a joint initiative of RWJBarnabas Health and Rutgers, The State University of New Jersey, and are used with permission of the Data Governance Council.

## Tables

**Supplemental Table S2.** Broader CCB-First (Primary) Cohort — Detailed Demographics by Treatment Arm.

| Variable | Gabapentin<br>(n=5,994) | Pregabalin<br>(n=1,147) | Total<br>(N=7,141) |
| --- | --- | --- | --- |
| Age, mean (SD) | 70.5 (11.7) | 69.7 (11.2) | 70.3 (11.6) |
| Age, median (IQR) | 71 (62–79) | 70 (62–78) | 71 (62–79) |
| Female, n (%) | 3,327 (55.5) | 640 (55.8) | 3,967 (55.6) |
| CKD at baseline, n (%) | 879 (14.7) | 151 (13.2) | 1,030 (14.4) |
| Diabetes, n (%) | 1,787 (29.8) | 335 (29.2) | 2,122 (29.7) |
| Heart failure, n (%) | 469 (7.8) | 65 (5.7) | 534 (7.5) |
| Stroke, n (%) | 267 (4.5) | 39 (3.4) | 306 (4.3) |
| Depression, n (%) | 382 (6.4) | 94 (8.2) | 476 (6.7) |
| Neuropathy, n (%) | 220 (3.7) | 49 (4.3) | 269 (3.8) |
| ACE inhibitor at baseline, n (%) | 866 (14.4) | 133 (11.6) | 999 (14.0) |
| ARB at baseline, n (%) | 1,699 (28.3) | 342 (29.8) | 2,041 (28.6) |
| Person-years of follow-up | 7,079 | 1,366 | 8,445 |
| Incident dementia events, n | 76 | 6 | 82 |
| Event rate per 100 person-years | 1.07 | 0.44 | 0.97 |

**Supplemental Table S1.** Established Dementia Cohort (Corrected Ascertainment): Incident Encephalopathy by CCB Status at Gabapentinoid Initiation.

| Outcome | DHP-CCB<br>(N=101) | No CCB<br>(N=145) | Non-DHP<br>CCB<br>(N=8)<sup>†</sup> | IPTW HR, DHP-CCB vs. No CCB<br>(95% CI) | P-value |
| --- | --- | --- | --- | --- | --- |
| Incident<br>encephalopathy | 8 (7.9%) | 15<br>(10.3%) | 0 events<sup>†</sup> | 0.73 (0.30–1.76) | 0.48 |

Established dementia cohort encephalopathy IPTW Cox: gabapentinoid initiators with prior dementia diagnosis (N=246 in the DHP-CCB vs. no-CCB analytic subset). Corrected HR for DHP-CCB vs. no-CCB exposure on incident encephalopathy: 0.73 (95% CI 0.30–1.76, p=0.48); 8 events of 101 DHP-CCB-exposed (7.9%); 15 events of 145 no-CCB (10.3%); 0 events of 8 non-DHP-CCB descriptive. The corrected null contrasts with the uncorrected analysis (HR 3.18, 95% CI 1.36–7.46), which was inflated by intravenous nicardipine prescriptions captured during inpatient hypertensive emergencies; the corrected analysis restricts to chronic outpatient DHP-CCB exposure. <sup>†</sup>The non-DHP-CCB arm (N=8, 0 events) is reported descriptively; insufficient for Cox estimation. Falls and dementia-medication-initiation outcomes reported in v13 are not shown pending re-analysis under the corrected ascertainment.

